# Data and code availability statements in systematic reviews of interventions were often missing or inaccurate: a content analysis

**DOI:** 10.1101/2021.12.06.21267383

**Authors:** Matthew J Page, Phi-Yen Nguyen, Daniel G Hamilton, Neal R Haddaway, Raju Kanukula, David Moher, Joanne E McKenzie

## Abstract

**Objectives:** To estimate the frequency of data and code availability statements in a random sample of systematic reviews with meta-analysis of aggregate data, summarise the content of the statements and investigate how often data and code files were shared.

**Methods:** We searched for systematic reviews with meta-analysis of aggregate data on the effects of a health, social, behavioural or educational intervention that were indexed in PubMed, Education Collection via ProQuest, Scopus via Elsevier, and Social Sciences Citation Index and Science Citation Index Expanded via Web of Science during a four-week period (between November 2^nd^ and December 2^nd^, 2020). Records were randomly sorted and screened independently by two authors until our target sample of 300 systematic reviews was reached. Two authors independently recorded whether a data or code availability statement (or both) appeared in each review and coded the content of the statements using an inductive approach.

**Results:** Of the 300 included systematic reviews with meta-analysis, 86 (29%) had a data availability statement and seven (2%) had both a data and code availability statement. In 12/93 (13%) data availability statements, authors stated that data files were available for download from the journal website or a data repository, which we verified as being true. While 39/93 (42%) authors stated data were available upon request, 37/93 (40%) implied that sharing of data files was not necessary or applicable to them, most often because “all data appear in the article” or “no datasets were generated or analysed”.

**Discussion:** Data and code availability statements appear infrequently in systematic review manuscripts. Authors who do provide a data availability statement often incorrectly imply that data sharing is not applicable to systematic reviews. Our results suggest the need for various interventions to increase data and code sharing by systematic reviewers.

What is new?

Key findings

- Data availability statements appeared in 31% of a random sample of 300 systematic reviews of interventions indexed during a four-week period in late 2020.
- 40% of authors who provided a data availability statement implied that sharing of data files was not necessary or applicable to them, most often because “all data appear in the article” or “no datasets were generated or analysed”.

What this adds to what is known?

- Data and code availability statements, which require authors to specify whether the data or code used in their study are available and if so, where it can be accessed, are required by some journals.
- It was unclear how often data and code availability statements appear in systematic review manuscripts, and what systematic reviewers typically write in their statements.

What is the implication and what should change now?

- Data and code availability statements should appear routinely in systematic review manuscripts. Doing so would conform to recommendations in the PRISMA 2020 statement.
- Journals should clarify in their instructions to authors that sharing of data files and, where relevant, analytic code, applies to systematic reviews.

## 1. BACKGROUND

Sharing files containing data, code (i.e. the sequence of commands used within a software package to manage and analyse data) and other materials that underlie a research project has many benefits. Sharing enables others to reuse the data to answer questions not considered by the original authors, check the data and code for errors, check how robust the results are across reasonable variations in analysis, and understand more about the study procedures and analyses than may be provided in the methods section of an article (1-3). To date, most of the research and commentary on sharing data and code in the health and medical field has been directed at studies which involve recruitment of participants (4-6). However, the benefits of sharing extend to research which does not require direct interaction with participants, such as systematic reviews with meta-analysis of aggregate study data (see Box 1 for examples of data generated by systematic reviewers which could be shared) (7). In addition to the benefits outlined above, sharing of reusable files containing data included in meta-analyses can facilitate updates of reviews, replication of reviews and inclusion of reviews in derivative products, such as overviews of systematic reviews or clinical practice guidelines, more rapidly than presenting a static table or figure in a manuscript can (8). Furthermore, sharing aggregate study data included in meta-analyses (e.g. summary statistics extracted from reports or retrieved from authors) does not require the same resources as sharing individual participant data does, meaning that the benefits of sharing are relatively easy to achieve.

To facilitate sharing of data and code, some journals require data and code availability statements in their manuscripts (9-11). These statements require authors to specify whether the data or code used in their study are available and if so, where it can be accessed (12). Authors can use such statements to indicate how FAIR (Findable, Accessible, Interoperable and Reusable) (13) the data and code files are, or indicate whether sharing was not applicable to them (e.g. a code sharing statement is not applicable to users of software without a command line interface, such as RevMan). Several studies have evaluated the frequency of data and code availability statements appearing in scientific manuscripts and found wide variation across disciplines (5, 14-18). For example, Serghiou et al. found that of 349 medical research articles indexed in PubMed from 2015 to 2018, 20% had a data availability statement and 1% had a code sharing statement (14); corresponding percentages were lower for psychology articles published from 2014-2017 (2% and 0.5%, respectively) (16). The content of data and code availability statements has been found to vary also (18-23). For example, analysis of 47593 data availability statements in articles published in PLOS ONE between March 2014 and May 2016 identified 10 types of statements (e.g. access to data is restricted, data are available in a repository, data are available upon request) (19). However, none of the previous studies specifically examined or analysed statements in systematic reviews.

It is unclear how often data and code availability statements appear in systematic review manuscripts, and what systematic reviewers typically write in their statements. Furthermore, there are a number of possible reasons why the prevalence of data sharing in systematic reviews may differ to that of primary studies. Systematic reviewers’ common requests for missing or incomplete data from primary studies might prime them to the importance of data sharing and influence them to share their own data. Alternatively, given the absence of discussion by journals and funders about sharing systematic review data and code, systematic reviewers may reasonably assume that the sharing of such material is not expected. Furthermore, recognition that the raw data to be shared is typically aggregate data which others could manually extract from tables and figures in a review manuscript might lead some systematic reviewers to assume sharing of files containing data and code is not applicable to them. To address this uncertainty, we sought to estimate the frequency of data and code availability statements in a random sample of systematic reviews with meta-analysis of aggregate data, summarise the content of the statements, and investigate how often data and code files were shared.

## 2. METHODS

This study arose as a sub-study of the REPRISE (REProducibility and Replicability In Syntheses of Evidence) project (24). As part of REPRISE, we evaluated completeness of reporting in a random sample of 300 systematic reviews of interventions indexed during a four-week period in 2020. The findings of the completeness of reporting evaluation will be reported elsewhere. Here, we provide a summary of our pre-specified methods used to identify systematic reviews, select reviews for inclusion, extract data from reviews and quantitatively analyse the extracted data; for full details see the published study protocol (24). In addition, we report the post-hoc methods we used to analyse the content of the data and code availability statements included in the systematic review reports. A protocol for these additional methods was not published or registered.

### 2.1. Eligibility criteria

We considered completed systematic reviews meeting the following criteria as eligible for inclusion:

- included randomized or non-randomized studies (or both) evaluating the effects of a health, social, behavioural or educational intervention on humans (studies needed to have compared one intervention with another or with no intervention);
- listed the full bibliographic reference for each study included in the review (we required this information for another component of the REPRISE project);
- presented the summary estimate for at least one pairwise meta-analysis of aggregate data, including at least two studies, irrespective of the chosen effect measure or its placement in the paper (i.e. the summary estimate could be presented in text, a table or a figure appearing in the main paper or supplementary files of the systematic review);
- written in English, given we did not have the resources to translate systematic reviews written in another language.

To be considered a “systematic review”, articles needed to have, at a minimum, clearly stated their review objective(s) or question(s); reported the source(s) (e.g. bibliographic databases) used to identify studies meeting the eligibility criteria, and reported conducting an assessment of the validity of the findings of the included studies, for example via an assessment of risk of bias or methodological quality. We did not exclude articles providing limited detail about the methods used. Systematic reviews with network meta-analyses were eligible if they included at least one pairwise (i.e. direct comparison) meta-analysis, while systematic reviews with only meta-analyses of individual participant data were excluded.

### 2.2. Search and selection of systematic reviews

We searched for eligible systematic reviews that were indexed during a four-week period (between November 2^nd^ and December 2^nd^, 2020) in PubMed, Education Collection via ProQuest, Scopus via Elsevier or Social Sciences Citation Index or Science Citation Index Expanded via Web of Science. We ran all searches on December 3^rd^, 2020. An information specialist created all search strategies (which are available in Appendix 1). All records yielded from each database were exported to Endnote vX8.2 software, and duplicate records were removed using automated record detection. We sorted the unique records in random order using the RAND() function in Microsoft Excel and uploaded the first 2000 records to Covidence (25) for screening. Two investigators (MJP and either PN or RK) independently and in duplicate screened the titles and abstracts of the 2000 records and retrieved any full text reports deemed (potentially) eligible. Two investigators (PN and either MJP or RK) then screened the full text reports until our target of 300 eligible systematic reviews was included. Any discrepancies in screening decisions were resolved via discussion or adjudication from the alternative investigator in the trio.

### 2.3. Sample size

The sample size of 300 was determined for the REPRISE study where we wished to estimate the percentage of reviews reporting a particular item (e.g. the search strategies for each database consulted) within a maximum margin of error (Wald type) of 6%, assuming a prevalence of 50%; for a prevalence of less (or greater) than 50%, the margin of error will be smaller. We deemed this margin of error to be generally sufficient such that our interpretation of the confidence limits would be consistent. For example, if we found the percentage of reviews reporting the search strategy for *each* database was 20% (95% CI 15% to 25%), our interpretation of the limits of the confidence interval would lead to the same conclusion, namely, that complete reporting of search strategies is not common.

### 2.4. Collection of data on systematic review characteristics

Two investigators (PN and either MJP, RK or ZA) independently and in duplicate collected data on characteristics of the 300 systematic reviews using a standardised and pilot-tested data collection form created in REDCap v10.6.12 (26). All discrepancies in data collected were resolved via discussion. The data collection form captured general characteristics of the review, such as country of corresponding author, type of intervention evaluated and funding source of the review, and items characterising the completeness of reporting of systematic reviews (items relevant to the current study are available in Appendix 2). We also recorded whether there was a data or code availability statement in the paper, and if there was, we recorded each statement verbatim. We defined a ‘data or code availability statement’ as either a journal-mandated, formalised statement about data/code availability that has its own dedicated section in the manuscript, or a statement appearing elsewhere in the manuscript (e.g. Methods or Results section) in which the authors state whether data/code files are available, and if so, how they can be accessed. For example, if authors stated at the end of the Methods section that “Data files are available upon request”, we classified the systematic review as having a data availability statement. Finally, we recorded whether particular types of data and code files were shared by the systematic reviewers, after checking the contents of every supplementary file for the review and following any links to repositories or personal websites the authors cited and confirming that the relevant files were located there.

### 2.5. Content analysis of data and code availability statements

We used an inductive approach (27) to categorise the content of each data and code availability statement. One investigator (MJP) read each statement and derived a code for each based on the meaning and content of the statement (e.g. the statement, “The data used to support the findings of this study are available from the corresponding author upon request” was coded as “Data available upon request”). As each subsequent statement was coded, existing codes were reviewed and revised, and new codes were added, where necessary. Once all statements were coded, the same investigator re-read each statement and code to ensure consistency in coding across the systematic reviews. A second investigator (PN) then classified each statement independently, using one of the codes populated by the first investigator or a new code they generated. Any discrepancies in the codes assigned to statements were resolved via discussion.

### 2.6. Quantitative data analysis

We summarised data on the characteristics of the systematic review as frequency and percentage for categorical items (e.g. country of corresponding author) and median and interquartile range for continuous items (e.g. number of included studies). We also calculated the frequency and percentage (with binomial exact 95% confidence intervals) of (i) systematic reviews which shared data and code files and (ii) systematic reviews with each category of data and code availability statement. Analyses were performed using the statistical package Stata, version 15 (28).

## 3. RESULTS

The searches yielded 8208 records, 6292 of which were unique (Figure 1). After screening titles and abstracts of the first 2000 randomly sorted records, we retrieved the full text of 603 articles. We needed to screen only the first 436 randomly sorted full text articles to reach our target of 300 eligible systematic reviews.

**Figure 1.**
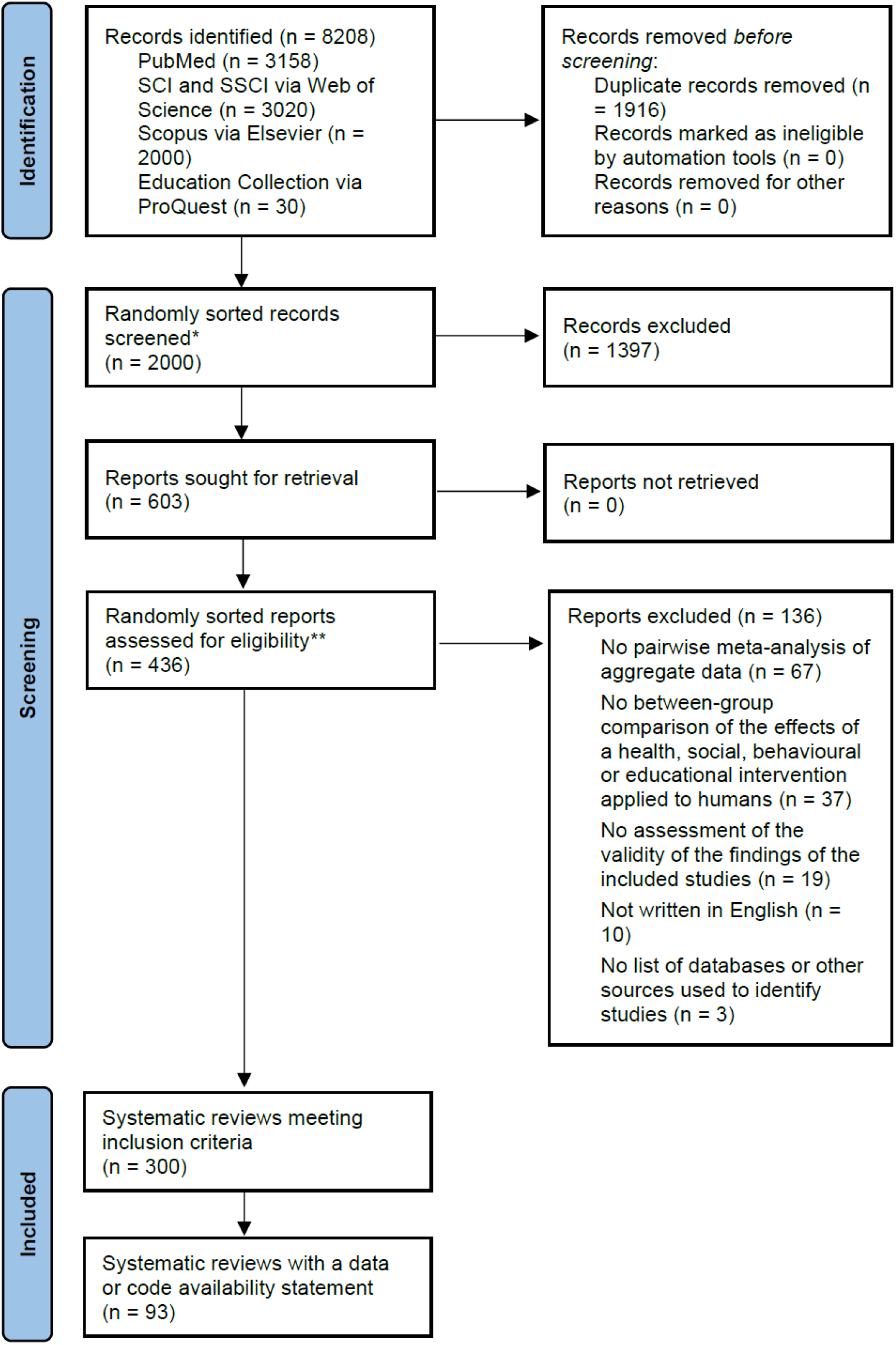
PRISMA 2020 flow diagram of identification, screening and inclusion of systematic reviews. *There were 6292 unique records after duplicates were removed, but we only needed to screen 2000 randomly sorted records to reach our target sample size of 300 reviews. **We only needed to screen 436 of the 603 full text reports retrieved to reach our target sample size of 300 reviews.

### 3.1. Characteristics of systematic reviews

Corresponding authors of the 300 systematic reviews were based in 45 countries, with most based in China (32%, 96/300), the United States of America (10%, 31/300) or the United Kingdom (8%, 24/300) (Table 1). Nearly all reviews (98%, 294/300) evaluated effects of a health intervention and 63% (184/294) of these were non-pharmacological interventions. The reviews addressed 23 different conditions or factors that influence health (classified using the 11^th^ revision of the International Classification of Diseases for Mortality and Morbidity statistics (29)), with diseases of the digestive system (12%, 36/300), endocrine, nutritional or metabolic diseases (12%, 36/300) and diseases of the musculoskeletal system (12%, 35/300) the most common. The most common type of funding source for reviews was a non-profit source (37%, 112/300). Most reviews were authored by individuals declaring no financial conflicts of interest (84%, 251/300), and were not registered nor had a protocol (58%, 174/300). The most common software packages used to conduct meta-analyses were RevMan (63%, 189/300), Stata (24%, 73/300) and R (11%, 33/300). The reviews included a median of 12 studies (interquartile range 8-21).

**Table 1.** Characteristics of included systematic reviews with meta-analysis (n=300)

| **Characteristic** | **Frequency (%)** |
| --- | --- |
| Country of corresponding author |  |
| China | 96 (32) |
| United States of America | 31 (10) |
| United Kingdom | 24 (8) |
| Iran | 15 (5) |
| Brazil | 14 (5) |
| Other | 120 (40) |
| Type of health condition addressed (ICD-11) |  |
| Diseases of the digestive system | 36 (12) |
| Endocrine, nutritional or metabolic diseases | 36 (12) |
| Diseases of the musculoskeletal system | 35 (12) |
| Diseases of the circulatory system | 30 (10) |
| Neoplasms | 29 (10) |
| Other health condition | 129 (43) |
| Not applicable | 5 (2) |
| Type of intervention addressed |  |
| Health | 294 (98) |
| Pharmacological | 103 (35) |
| Non-pharmacological | 184 (63) |
| Both | 7 (2) |
| Social | 5 (2) |
| Behavioural | 28 (9) |
| Educational | 4 (1) |
| Number of included studies<sup>1</sup> | 12 (8-21) |
| Source of funding |  |
| Non-profit | 112 (37) |
| For-profit | 3 (1) |
| Both non-profit and for-profit | 3 (1) |
| Authors specified there was no funding | 97 (32) |
| Not reported | 85 (28) |
| Financial conflicts of interest of systematic reviewers |  |
| At least one systematic reviewer reported a financial conflict of interest | 30 (10) |
| All systematic reviewers stated they had no financial conflicts of interest | 251 (84) |
| No disclosure statement | 19 (6) |
| Systematic review protocol or registration mentioned |  |
| Both a protocol and registration record cited | 1 (1) |
| Only a protocol cited | 13 (4) |
| Only a registration record cited | 112 (37) |
| Neither cited | 174 (58) |
| Software used to conduct meta-analysis<sup>2</sup> |  |
| RevMan | 189 (63) |
| Stata | 73 (24) |
| R | 33 (11) |
| Comprehensive Meta-Analysis | 27 (9) |
| Other | 18 (6) |
| Not reported | 3 (1) |

<sup>1</sup>Data are given as median (interquartile range). <sup>2</sup>Percentages do not sum to 100 as more than one type of software might have been used.

### 3.2. Content of data and code availability statements

Of the 300 reviews, seven (2%) had both a data and code availability statement, 86 (29%) had only a data availability statement and none had only a code availability statement. In 13% (12/93) of the data availability statements, authors stated that data files were available for download from the journal website or a data repository (Table 2). In 42% (39/93) of cases, authors stated data were available upon request, whereas in 40% (37/93), authors implied that sharing of data files was not necessary or applicable to them. Specific reasons provided were that all data are in the article or supplementary file (despite no data files being made available) (23%, 21/93), that no datasets were generated or analysed (10%, 9/93) or the data are from the published literature (8%, 7/93). The remaining five data availability statements were either conflicting (e.g. stated that no datasets were generated yet that the datasets generated were available upon request), indicated that data will be made available in future, or that data were not available. Of the seven code availability statements, authors stated either “not applicable” (n=4), that code was available upon request (n=1), or that code was publicly available and where it could be accessed (n=2).

**Table 2.** Classifications and illustrative quotes of data and code availability statements in systematic reviews with meta-analysis.

| Classification | Illustrative quote | Freq. | % (95% CI) |
| --- | --- | --- | --- |
| ***Data availability statements (n=93)*** |  |  |  |
| Data are available upon request | “The data used to support the findings of this study are available from the corresponding author upon request.” | 39 | 42% (32%-53%) |
| All data are in the article and supplementary files | “All relevant data are within the manuscript and its supporting information files.” | 21 | 23% (15%-32%) |
| Data files are available for download from the journal website or a data repository | “The dataset was openly provided [link provided].” | 12 | 13% (7%-21%) |
| Not applicable as no datasets were generated or analysed | “Data sharing is not applicable to this article as no datasets were generated or analyzed during the current study.” | 9 | 10% (5%-18%) |
| Not applicable because the data are from the published literature | “Data sharing is not applicable as the data are secondary data drawn from already published literature.” | 7 | 8% (3%-15%) |
| Conflicting statement | “Data sharing not applicable to this article as no datasets were generated or analyzed during the current study. The datasets generated during and/or analyzed during the current study are available from the corresponding author on reasonable request.” | 3 | 3% (1%-9%) |
| Data will be made available in future | “Following acceptance and before publication, all data will be provided on a public repository.” | 1 | 1% (0%-6%) |
| Data are not available | “No data are available.” | 1 | 1% (0%-6%) |
| ***Code availability statements (n=7)*** |  |  |  |
| Not applicable | “Code availability: Not applicable.” | 4 | 57% (18%-90%) |
| Code is available in a public, open access repository or supplementary file | “...codes used for network meta-analysis can be found in data.bris repository [link provided].” | 2 | 29% (4%-71%) |
| Code is available upon request | “The full data set and statistical code is available from [corresponding author name].” | 1 | 14% (0%-58%) |

The 93 systematic reviews with a data availability statement were published in 59 journals. There were 10 journals publishing more than one of these systematic reviews, and the content of the data availability statements was identical within the systematic reviews published in two of these journals: *Cochrane Database of Systematic Reviews* (8 systematic reviews) and *PLOS ONE* (6 systematic reviews). Authors of all the Cochrane Reviews stated that data files were available for download from the journal website whereas authors of all the *PLOS ONE* reviews stated that all data are in the article or supplementary files.

### 3.3. Frequency and type of systematic review data and code files shared

Sharing of systematic review data files and code was rare in our sample (Table 3). In 4% (12/300) of reviews, authors shared the file(s) containing data used in all analyses (e.g. Microsoft Excel or CSV spreadsheet, or rm5 file containing all study effect estimates included in meta-analyses). Fewer authors shared the file(s) containing (unprocessed) data extracted from included studies (3%, 9/300), code used to generate results (e.g. the sequence of commands used within R or Stata to manage and analyse data) (0.7%, 2/300) or metadata which describes the contents of the shared file(s) to aid interpretation and reuse (e.g. a file with complete descriptions of variable names, or README files describing each file shared) (0.3%, 1/300). Of the 20 systematic reviews with at least one type of file shared, at least one file was uploaded as a supplement on the journal website in 20 (100%) cases, at least one file was uploaded to a general purpose open-access repository in one (5%) case, and at least one file was uploaded to an institutional repository in one (5%) case. Furthermore, for only two (10%) of these 20 reviews was a permanent identifier (e.g. DOI) or license outlining the terms of use (e.g. CC BY) assigned to the shared file(s).

**Table 3.** Frequency and type of systematic review data and code files shared (n=300)

| Item | Freq. | % (95% CI) |
| --- | --- | --- |
| *Type of material shared* |  |  |
| Template data collection form(s) | 0 | - |
| File(s) containing (unprocessed) data extracted from included studies | 9 | 3% (1%-6%) |
| File(s) indicating any necessary data conversions performed | 1 | 0.3% (0%-2%) |
| File(s) containing data used in all analyses (e.g. Microsoft Excel or CSV spreadsheet, or RevMan file containing all study effect estimates included in meta-analyses) | 12 | 4% (2%-7%) |
| Analytic code used to generate results (i.e. the sequence of commands used within a software package to manage and analyse data) | 2 | 0.7% (0%-2%) |
| Metadata which describes the contents of the shared file(s) to aid interpretation and reuse (e.g. a file with complete descriptions of variable names, or README files describing each file shared) | 1 | 0.3% (0%-2%) |
| *Location where any of the above materials was shared* |  |  |
| Journal website (i.e. as a supplementary file) | 20* | 100% (83%-100%) |
| General purpose open-access repository (e.g. OSF, Zenodo) | 1* | 5% (0.1%-25%) |
| Institutional repository | 1* | 5% (0.1%-25%) |
| *Persistent identifier and licensing for any of the above material shared* |  |  |
| Persistent identifier (e.g. DOI) assigned to the shared file(s) | 2* | 10% (1%-32%) |
| License outlining the terms of use (e.g. CC BY) assigned to the shared file(s) | 2* | 10% (1%-32%) |

\*Denominator is 20 systematic reviews in which at least one type of material was shared

## 4. DISCUSSION

Based on our analysis of 300 systematic reviews of health interventions indexed in late 2020, sharing of data files and code or publicly declaring an intention to do so is rarely done by systematic reviewers. Actual sharing of data or code files was observed in only 12 reviews and a statement that data were available upon request appeared in only 39 reviews. The latter type of statement does not necessarily guarantee data will be made available, as authors can (and often do) choose to decline or ignore requests (23). Furthermore, several authors incorrectly implied that sharing of data files is not applicable to systematic reviews, with the reasons for this perspective varying.

To our knowledge, this is the first study to explore data and code availability statements appearing in systematic reviews, so we can only compare our findings to evaluations of other types of research article. The percentage of articles with data and code availability statements observed in our study (31% and 2%, respectively) is similar to that observed in other studies examining a sample of health research articles, such as the evaluation conducted by Serghiou et al. (20% and 1%, respectively) (14), Wallach et al. (18% and 0%, respectively) (17) and Naudet et al. (25% with a data availability statement) (5). Also, the percentage of data availability statements naming a publicly accessible location where the data are available (13% in our study) was similar to that observed by Federer et al. (19) in their analysis of articles published in PLOS ONE (15%) and by Colavizza et al. (20) in their analysis of articles published in BMC journals (12%). These similarities suggest that systematic reviewers share the same awareness of and attitudes towards data and code sharing as authors of other study designs.

Some of the provided reasons for not sharing systematic review data files are misguided. The claim that no datasets were generated or analysed – and hence there was no data to share – is inaccurate given meta-analysis, which involves the collection and analysis of data from a set of studies, was undertaken in all reviews included in this study. Other types of data generated by systematic reviewers include risk of bias assessments, certainty of evidence assessments, and classifications of study design features (see Box 1). Also, the assertion that systematic review data need not be shared because it is “secondary data drawn from the published literature” is questionable; researchers should consider sharing systematic review data regardless of from where it has been drawn. Data from primary studies often requires some manipulation (e.g. imputing missing summary statistics, converting results to a standardised effect measure, correcting mistakes found in the primary publications, including unpublished statistics obtained from primary study authors [e.g. data for particular subgroups]) before its inclusion in meta-analysis, so it is insufficient to simply point readers to the study papers. Furthermore, while it is often true that all data for a meta-analysis are “in the systematic review article” when authors present a forest plot or table with relevant study results, sharing an editable file (e.g. CSV, RevMan) containing such data would facilitate its reuse, as it would prevent the need for others to manually extract data from the review, thus saving time and reducing data extraction errors.

Several interventions could be employed to increase data sharing and ensure data availability statements in systematic reviews are appropriate. For example, educational programs on data sharing, and open science more broadly, could be offered to undergraduate and graduate students working on systematic reviews. Journals could mandate (or at a minimum encourage) systematic reviewers to share their data and prevent publication of any misleading data availability statements. We read the instructions to authors of all journals publishing a systematic review with a data availability statement in our sample, and in none was there a recommendation to write “not applicable” if submitting a systematic review. However, none of the journals singled out systematic reviews as a type of research for which data and code sharing would be beneficial, so more explicit encouragement might be necessary. Furthermore, journals could provide clearer instructions to authors about which data availability statements may or may not apply in the context of systematic reviews. For example, authors of systematic reviews could be advised to avoid statements such as “No datasets were generated or analysed during the current study”. In addition, funders could make it an expectation that the data for systematic reviews they fund be shared in accordance with the FAIR principles. Finally, academic institutions could reward systematic reviewers who make their review data and code publicly accessible, in keeping with the Hong Kong Principles for assessing researchers (30).

The rarity of code availability statements observed in our sample of systematic reviews may be explained by several factors. Authors of only 112 (37%) of the 300 systematic reviews used statistical software with either command line interfaces or graphical user interfaces that allow users to extract the code that was run (e.g. R, Stata, SAS, SPSS). Therefore, users of alternative software (e.g. RevMan) might have considered it unnecessary to include a code availability statement, given no such code was able to be shared. In addition, journals are more likely to require or encourage inclusion of a data availability statement than a code availability statement (31), so without prompting, systematic reviewers might not have even considered the value of including a code availability statement. Furthermore, some systematic reviewers who use software which enables code sharing might not know how to share their code, feel uncomfortable with sharing the code due to concerns that errors might be detected in it or embarrassed about how it was written. As part of the REPRISE project (24), we plan to survey authors of systematic reviews to explore their views on sharing systematic review data and code, which should help elucidate this issue further.

Our findings should be considered in relation to the strengths and limitations of the study. Two authors independently screened all records, collected data from all reviews, and coded all data and code availability statements, to minimise the risk of errors in these steps. Our definition of ‘data and code availability statements’ was broad enough to capture systematic reviews in which authors were required to complete a journal-mandated data/code availability statement or indicated somewhere in text whether data/code were available. Also, by not limiting our focus to systematic reviews published in specific journals, our findings can be generalised to a larger population of reviews. However, a limitation of our study is that we only examined the PDF and online version of each review manuscript to locate data and code availability statements and links to shared files. It is possible that authors made such files available (e.g. deposited them in a repository or on a personal/institutional website) after the article was published. Given we did not perform a search of data repositories for each authors’ data/code files, we might have underestimated the true frequency of data/code sharing. Also, the coding framework we developed in our content analysis of the data and code availability statements might have been influenced by our exposure to literature on this topic and our prior experiences with writing such statements for our own manuscripts. Therefore, it is possible that other researchers might have classified the statements differently. We have made our dataset freely available for scrutiny by others (see https://osf.io/ya4hp/).

We believe data and code availability statements should appear routinely in systematic review manuscripts. Doing so would conform to recommendations in the PRISMA 2020 statement (published in March 2021, after the reviews we evaluated were published), which advises authors to “Report which of the following are publicly available and where they can be found: template data collection forms; data extracted from included studies; data used for all analyses; analytic code; any other materials used in the review” (32, 33). Furthermore, we believe there are only a few situations where authors are unable to share systematic review data files, such as when the review authors are custodians rather than owners of individual participant data; no such restriction applies to systematic reviews with meta-analysis of aggregate data. Authors who used software that does not allow them to extract the code that was run should still include a code availability statement, which specifies why they are unable to share their code. For guidance on how to share systematic review data, code and other materials, see Box 2; we also recommend authors consult the FAIR (Findable, Accessible, Interoperable, Reusable) guiding principles (13). Adopting such principles should ultimately enhance the reproducibility of systematic reviews and their value to users.

## Data Availability

The data and analytic code for this study are available on the Open Science Framework at https://osf.io/ya4hp/ (DOI: 10.17605/OSF.IO/YA4HP).

## Acknowledgments

We thank Steve McDonald for developing the search strategy. We thank Zainab Alqaidoom for extracting data from 50 of the 300 included systematic reviews.

## Authors’ contributions

All authors declare to meet the ICMJE conditions for authorship. MJP and JM conceived the study design. MJP, PN and RK screened articles for eligibility and collected data. MJP and PN coded data availability statements. MJP analysed the data. MJP wrote the first draft of the article. All authors contributed to revisions of the article. All authors approved the final version of the submitted article.

## Box 1.

**Examples of data generated by systematic reviews which could be shared via reusable files**

- Numeric values extracted from study reports to calculate study effect estimates and measures of precision
- Calculated study effect estimates and their variances
- Results of meta-analyses conducted, such as the summary effect estimate, its precision and estimates of heterogeneity and inconsistency
- Risk of bias assessments for each study
- Certainty of evidence assessments for each outcome
- Classifications of study design features
- Classifications of the interventions across the studies using a standardised system

## Box 2.

**Recommendations for data and code sharing for systematic reviews**

*What to share*:

- Data such as that referred to in Box 1.
- Analytic code used to generate results.
- Metadata (such as README files describing each data file shared).

*Where to share:*

- In one of the general, domain-specific or local institutional repositories listed on https://www.re3data.org/. Commonly used general repositories include the Open Science Framework, Dryad and figshare. The Systematic Review Data Repository (SRDR) is an example of a repository for sharing materials specific to systematic reviews.

*How to share:*

- Ideally each file shared should be available in an accessible format (such as a CSV or .txt file). If a specific software file is shared (e.g. a .rm5 file from Review Manager – Cochrane’s software for preparing and maintaining Cochrane reviews (34)), details of what software package is required to use the data should be provided in the metadata file(s) (e.g. README file).
- Assign a persistent identifier (e.g. DOI) and license outlining the terms of use (e.g. CC BY) to each file shared.

