## Appendix for "Data and code availability statements in systematic reviews of interventions were often missing or inaccurate: a content analysis"

**Appendix 1: Search strategies**

Searches run on Thursday 3 December 2020

**PubMed**
(meta-analysis[PT] OR meta-analysis[TI] OR systematic[sb]) AND 2020/11/02:2020/12/02[EDAT]

**Science Citation Index Expanded (SCI-EXPANDED) and Social Sciences Citation Index (SSCI) via Web of Science**
(TI=meta-analysis OR AB=meta-analysis OR TS=meta-analysis OR TI="systematic review" OR AB="systematic review" OR TS="systematic review") AND **LANGUAGE**: (English) AND **DOCUMENT TYPES**: (Article OR Review)
*Indexes=SCI-EXPANDED, SSCI; Timespan=Last 4 weeks*

**Scopus via Elsevier**
TITLE("meta-analysis" OR "systematic review") AND ORIG-LOAD-DATE > 1604275200 AND ORIG-LOAD-DATE < 1606867200 AND (LIMIT-TO(DOCTYPE, "ar") OR LIMIT-TO(DOCTYPE, "re"))

**Education Collection via ProQuest (added in 30 last days)**
MAINSUBJECT.EXACT.EXPLODE("Meta Analysis") OR ab(meta-analysis OR systematic review) OR ti(meta-analysis OR systematic review)

**Appendix 2. Data collection form**

| Item | Response option |
|---|---|
| Specify the title of the systematic review: | Free text |
| Specify the journal: | Free text |
| Specify the country of the corresponding author of the systematic review: | Free text |
| What was the source of funding for the systematic review? | o Non-profit (e.g. government, university/hospital/research institute, charitable foundation)<br>o For-profit (e.g. pharmaceutical company)<br>o Both non-profit and for-profit<br>o Unclear if the funder is for-profit or non-profit (please specify funder): ____________________<br>o Authors specified there was no funding for the systematic review<br>o Not reported |
| Did any of the systematic reviewers disclose financial conflicts of interest? | o Conflict of interest present (i.e. at least one systematic reviewer reported a financial conflict of interest of any type)<br>o No conflict of interest (i.e. all systematic reviewers stated they had no financial conflicts of interest)<br>o Missing (i.e. no disclosure statement) |
| Specify the total number of studies included in the systematic review: | Numeric |
| What is the broad ICD-11 category investigated in this systematic review? | o Not applicable<br>o 01 Certain infectious or parasitic diseases<br>o 02 Neoplasms<br>o 03 Diseases of the blood or blood-forming organs<br>o 04 Diseases of the immune system<br>o 05 Endocrine, nutritional or metabolic diseases<br>o 06 Mental, behavioural or neurodevelopmental disorders<br>o 07 Sleep-wake disorders<br>o 08 Diseases of the nervous system<br>o 09 Diseases of the visual system<br>o 10 Diseases of the ear or mastoid process<br>o 11 Diseases of the circulatory system<br>o 12 Diseases of the respiratory system<br>o 13 Diseases of the digestive system<br>o 14 Diseases of the skin<br>o 15 Diseases of the musculoskeletal system or connective tissue<br>o 16 Diseases of the genitourinary system |

| Item | Response option |
|---|---|
| | ○ 17 Conditions related to sexual health<br>○ 18 Pregnancy, childbirth or the puerperium<br>○ 19 Certain conditions originating in the perinatal period<br>○ 20 Developmental anomalies<br>○ 21 Symptoms, signs or clinical findings, not elsewhere classified<br>○ 22 Injury, poisoning or certain other consequences of external causes<br>○ 23 External causes of morbidity or mortality<br>○ 24 Factors influencing health status or contact with health services |
| Specify the type of intervention(s) indicated in the review objective(s) or question(s) (e.g. inhaled corticosteroids, provision of charity or welfare, use of bystander programs, reduction in class size) [Select all that apply]: | ○ Health (i.e. any intervention designed to improve "health" defined as a state of complete physical, mental and social well-being and not merely the absence of disease or infirmity)<br>○ Behavioural (i.e. any intervention designed to increase useful behaviours and reduce or eliminate harmful behaviours, such as modelling, prompting, reinforcement, or shaping behaviours)<br>○ Educational (i.e. any intervention designed to provide students with the support needed to acquire the skills being taught by an educational system)<br>○ Social (i.e. any intervention of a government or an organization in social affairs, such as provision of charity or social welfare as a means to alleviate social and economic problems of people facing financial difficulties; provision of safety regulations for employment and products; delivery of food aid, food bank or recovery missions to regions or countries negatively affected by an event; securing workers' rights through gradualism in institutions, private education and unions) |
| Is/are the intervention intervention(s) indicated in the review objective(s) or question(s) pharmacological or non-pharmacological? | ○ Pharmacological<br>○ Non-pharmacological (i.e. any intervention that does not involve drugs, e.g. device, vitamin, diet, psychotherapy, physical therapy, welfare, organization change, education)<br>○ Both pharmacological and non-pharmacological |
| Is a protocol or registration record for the systematic review cited within the review? | ○ Both a protocol and registration record are cited<br>○ Only a protocol is cited |

| Item | Response option |
|---|---|
| | ○ Only a registration record is cited<br>○ Neither are cited |
| Which statistical software was used to perform meta-analyses [Select all that apply]? | ○ R<br>○ Stata<br>○ RevMan<br>○ Comprehensive Meta-Analysis (CMA)<br>○ SAS<br>○ SPSS<br>○ Other (please specify)<br>○ Not reported |
| Is there a data or code availability statement in the paper? | ○ Yes<br>○ No |
| If yes to the previous question, copy and paste the data and code statement(s) verbatim: | Free text |
| Have the authors made any of the following publicly accessible, either as a supplementary file or file uploaded to a general-purpose repository (e.g. Open Science Framework, Zenodo, GitHub) or institutional repository? [Select all that apply. Only select each if you are able to locate the relevant material, after searching all supplementary materials or links to shared materials] | ○ Template data collection form(s)<br>○ File(s) containing (unprocessed) data extracted from included studies<br>○ File(s) indicating any necessary data conversions performed<br>○ File(s) containing data used in all analyses (e.g. Microsoft Excel or CSV spreadsheet, or RevMan file containing all study effect estimates included in meta-analyses)<br>○ Analytic code used to generate results (i.e. the sequence of commands used within a software package to manage and analyse data)<br>○ File(s) containing citations of all records that were screened and excluded<br>○ Metadata which describes the contents of the shared file(s) to aid interpretation and reuse (e.g. a file with complete descriptions of variable names, or README files describing each file shared) |
| If yes to any of the materials selected, how was the material shared publicly? [Select all that apply] | ○ Uploaded as a supplementary file on the journal website<br>○ Uploaded to a general-purpose open-access repository (e.g. Open Science Framework, Zenodo, GitHub) (please specify)<br>○ Uploaded to an institutional repository<br>○ Uploaded to a personal website<br>○ Other (please specify) |
| If yes to any of the materials selected, are any of the data file(s) associated with a persistent identifier (e.g. DOI)? | ○ Yes<br>○ No<br>○ Unsure |

| Item | Response option |
|---|---|
| If yes to any of the materials selected, was a license applied to any of the data file(s) (e.g. CC BY, CC BY-NC)? | ○ Yes<br>○ No<br>○ Unsure |
